# BEYOND THE SURFACE: VALIDATING THE ANTHROPOMETRIC-BIOCHEMICAL LINK IN CHILDHOOD MALNUTRITION IN SOKOTO STATE, NIGERIA

**DOI:** 10.64898/2026.03.18.26348747

**Authors:** Ibrahim Ashafura Musa, Kabir M Y

## Abstract

Anthropometric indicators such as stunting, wasting, and underweight are the primary tools for diagnosing childhood malnutrition in resource-limited settings. However, the extent to which these visible signs reflect underlying biochemical derangements remains inadequately characterized. This study aimed to examine the associations between anthropometric indicators of malnutrition and key nutritional biomarkers among under-five children in Sokoto State, Nigeria. A community-based cross-sectional study was conducted among 150 mother-child pairs attending Primary Health Centres. Anthropometric measurements (weight, height/length, MUAC) were collected and used to classify children by nutritional status (stunting, wasting, underweight). Venous blood samples were analyzed for Prealbumin, C-Reactive Protein (CRP), Serum Retinol (Vitamin A), Hemoglobin, Serum Albumin, and Serum Zinc. Statistical analyses included independent t-tests, Pearson’s correlation, chi-square tests for trend, and binary logistic regression. Children classified as malnourished by MUAC (<12.5 cm) had significantly worse biomarker profiles than their well-nourished counterparts (p < 0.001 for all biomarkers). A strong, dose-response relationship was observed: the prevalence of vitamin A deficiency increased from 72.3% in normal children to 95.1% in MAM and 100% in SAM; zinc deficiency from 38.3% to 78.7% to 95.2%; and anemia from 46.8% to 85.2% to 97.6%. MUAC showed the strongest correlations with biomarkers (r = 0.56-0.71, p < 0.01). Critically, 72.3% of anthropometrically “normal” children had vitamin A deficiency, and 38.3% had three or more concurrent deficiencies. Logistic regression revealed that children with SAM had 18-43 times higher odds of biochemical deficiencies compared to normal children Anthropometric status strongly predicts biochemical depletion, validating the use of MUAC for identifying children at highest risk. However, the high burden of “hidden hunger” among anthropometrically normal children reveals a critical limitation of sole reliance on anthropometry. These findings argue for integrated assessment approaches and multi-micronutrient interventions to address the full spectrum of malnutrition, from visible wasting to invisible biochemical deficiencies.

## 1. INTRODUCTION

Childhood malnutrition remains a persistent public health challenge in low- and middle-income countries, with Nigeria bearing one of the highest burdens globally (Black et al., 2013; National Population Commission [NPC] & ICF, 2019). In Sokoto State, located in Nigeria’s Northwest region, previous research has documented alarming rates of stunting, wasting, and underweight among children under five years. However, anthropometric indicators while essential for screening and diagnosis in resource-limited settings provide only an indirect measure of nutritional status. They capture the visible consequences of malnutrition but may not fully reflect the underlying physiological reality (Gibson, 2005).

The concept of “hidden hunger” chronic deficiencies in essential vitamins and minerals that may not manifest in visible growth failure has gained increasing recognition in global nutrition discourse (Muthayya et al., 2013). Children who appear well-nourished by simple arm circumference measurement may nonetheless be severely deficient in micronutrients critical for immune function, cognitive development, and long-term health (Bailey et al., 2015). Conversely, children identified as wasted or stunted through anthropometry are presumed to have significant nutritional deficits, but the precise biochemical nature and severity of these deficits remain poorly quantified in many high-burden settings (Raiten et al., 2015).

Understanding the relationship between anthropometric indicators and biochemical markers is critical for several reasons. First, it validates the use of anthropometry as a proxy for biochemical status, confirming that visible signs of malnutrition indeed reflect underlying physiological depletion. Second, it elucidates the specific biochemical pathways through which malnutrition manifests, potentially identifying targets for intervention. Third, it quantifies the extent to which anthropometric deficits correspond to biochemical derangements, informing clinical decision-making and program design. Fourth, it reveals the limitations of anthropometry alone, particularly its inability to detect “hidden hunger” in children who appear well-nourished (Thurnham et al., 2021).

This study was designed to address these critical evidence gaps. The primary objective was to examine the associations between anthropometric indicators of malnutrition (stunting, wasting, underweight, MUAC) and the levels of key nutritional biomarkers (Prealbumin, CRP, Serum Retinol, Hemoglobin, Albumin, Zinc) among under-five children in Sokoto State. By integrating anthropometric and biochemical data, this study aims to provide a more comprehensive understanding of the malnutrition syndrome in this population and to generate evidence for more effective, integrated intervention strategies (Raiten et al., 2015).

## 2. Methodology

### 2.1 Study Design and Setting

This study employed a community-based, cross-sectional design. It was conducted in selected Primary Health Centres (PHCs) across Local Government Areas (LGAs) in Sokoto State, Northwest Nigeria, between September, 2024 and September, 2025. Sokoto State is characterized by high poverty rates, chronic food insecurity, and limited access to healthcare and clean water.

### 2.2 Study Population and Sampling

The study population comprised children aged 0 to 59 months and their mothers/caregivers who presented at the selected PHCs for routine child welfare services. A total of 150 mother-child pairs were enrolled using a multi-stage sampling technique. Children with physical deformities that could impede accurate anthropometric measurement, those who were critically ill and requiring immediate emergency care, or those with known chronic illnesses (e.g., sickle cell disease) were excluded from the study.

### 2.3 Data Collection

Data were collected through face-to-face interviews with mothers or caregivers using a semi-structured, pre-tested questionnaire. Anthropometric measurements were conducted using standardized techniques and calibrated equipment: weight was measured to the nearest 0.1 kg using a Seca electronic scale with children wearing minimal clothing; recumbent length was measured for children under 24 months using a calibrated infantometer, while standing height was measured for older children using a portable stadiometer, both recorded to the nearest 0.1 cm; and mid-upper arm circumference (MUAC) was measured at the midpoint of the left upper arm using a non-stretchable MUAC tape, recorded to the nearest 0.1 cm. Venous blood samples (approximately 3 to 5 mL) were collected from each child by a trained phlebotomist under aseptic conditions and transported in a cold chain to a diagnostic laboratory for analysis. Assays included prealbumin (transthyretin) and C-reactive protein (CRP) by high-sensitivity immunoturbidimetry; serum retinol (vitamin A) by high-performance liquid chromatography (HPLC); hemoglobin by hematology analyzer; serum albumin by bromocresol green (BCG) dye-binding method; and serum zinc by atomic absorption spectrophotometry.

### 2.4 Data Analysis

Data were entered and analyzed using IBM SPSS Statistics version 26, with anthropometric indices including Weight-for-Age Z-score (WAZ), Height-for-Age Z-score (HAZ), and Weight-for-Height Z-score (WHZ) calculated using WHO Anthro software (version 3.2.2) and nutritional status classified according to WHO Child Growth Standards (WHO, 2006). MUAC was categorized using standard cut-offs: normal (>12.5 cm), moderate acute malnutrition (MAM: 11.5–12.5 cm), and severe acute malnutrition (SAM: <11.5 cm), while biomarker deficiencies were defined using internationally recognized thresholds (WHO, 2011; IZiNCG, 2004; Ingenbleek & Young, 1994). Three complementary analytical approaches were employed: comparison of means using independent t-tests and ANOVA to assess differences in biomarker levels across nutritional categories; correlation analysis using Pearson’s coefficients to examine relationships between anthropometric indices and biomarker concentrations; prevalence analysis through cross-tabulation with chi-square tests for trend to determine biochemical deficiency rates within each anthropometric group; and binary logistic regression to quantify the predictive value of MUAC category for identifying children with biochemical deficiencies. Statistical significance was set at p < 0.05, and all tests were two-tailed.

### 2.5 Ethical Considerations

This study adhered to the ethical principles of the Declaration of Helsinki. Ethical approval was obtained from the Health Research Ethics Committee of the Sokoto State Ministry of Health (Ref: SKHREC/059/2022). Permission was also secured from the SSPHCDA and local government authorities.

## 3. Results and Discussions

### 3.1 Comparison of Mean Biomarker Levels by MUAC Classification

Independent samples t-tests revealed consistently and significantly lower levels of all nutritional biomarkers among children classified as malnourished by MUAC (<12.5 cm, n=103) compared to their well-nourished counterparts (>12.5 cm, n=47). These differences were highly statistically significant (p < 0.001) for all biomarkers assessed (Table 1).

**Table 1:** Comparison of Mean Biomarker Levels by Nutritional Status (MUAC Classification).

| Biomarker | Nutritional Status (MUAC) | N | Mean ± SD | Mean Difference | t-statistic | p-value | 95% CI of Difference |
| --- | --- | --- | --- | --- | --- | --- | --- |
| Prealbumin (mg/dL) | Malnourished (<12.5 cm) | 103 | 5.8 ± 1.7 | 2.8 | 6.94 | <0.001 | 2.0 – 3.6 |
| | Normal (>12.5 cm) | 47 | 8.6 ± 2.3 | | | | |
| CRP (mg/L) | Malnourished (<12.5 cm) | 103 | 6.7 ± 2.4 | -2.4 | -5.12 | <0.001 | -3.3 – -1.5 |
| | Normal (>12.5 cm) | 47 | 4.3 ± 1.9 | | | | |
| Serum Retinol (μmol/L) | Malnourished (<12.5 cm) | 103 | 0.52 ± 0.16 | 0.28 | 7.45 | <0.001 | 0.20 – 0.36 |
| | Normal (>12.5 cm) | 47 | 0.80 ± 0.22 | | | | |
| Hemoglobin (g/dL) | Malnourished (<12.5 cm) | 103 | 8.6 ± 1.0 | 1.9 | 8.21 | <0.001 | 1.4 – 2.4 |
| | Normal (>12.5 cm) | 47 | 10.5 ± 1.2 | | | | |
| Serum Albumin (g/dL) | Malnourished (<12.5 cm) | 103 | 2.8 ± 0.4 | 0.9 | 7.68 | <0.001 | 0.7 – 1.1 |
| | Normal (>12.5 cm) | 47 | 3.7 ± 0.5 | | | | |
| Serum Zinc (μg/dL) | Malnourished (<12.5 cm) | 103 | 57.3 ± 6.8 | 11.2 | 8.34 | <0.001 | 8.5 – 13.9 |
| | Normal (>12.5 cm) | 47 | 68.5 ± 7.9 | | | | |

Source: SPSS

Malnourished children also had significantly higher levels of the inflammatory marker CRP (6.7 vs 4.3 mg/L, p < 0.001), indicating that acute malnutrition is accompanied by systemic inflammation.

### 3.2 Comparison of Mean Biomarker Levels by Stunting Status

Stunted children (HAZ < −2SD, n=83) had significantly worse biomarker profiles compared to non-stunted children (HAZ ≥ −2SD, n=67) across all parameters, confirming that chronic linear growth failure is underlain by chronic nutritional deficiencies (Table 2).

**Table 2:** Comparison of Mean Biomarker Levels by Stunting Status (Height-for-Age Z-score).

| **Biomarker** | **Stunting Status** | **N** | **Mean ± SD** | **Mean Difference** | **t-statistic** | **p-value** |
| --- | --- | --- | --- | --- | --- | --- |
| Prealbumin (mg/dL) | Stunted (HAZ < -2SD) | 83 | 6.0 ± 1.9 | 1.7 | 4.23 | <0.001 |
| | Not Stunted (HAZ ≥ -2SD) | 67 | 7.7 ± 2.3 | | | |
| CRP (mg/L) | Stunted (HAZ < -2SD) | 83 | 6.3 ± 2.5 | -1.1 | -2.89 | 0.004 |
| | Not Stunted (HAZ ≥ -2SD) | 67 | 5.2 ± 2.1 | | | |
| Serum Retinol (μmol/L) | Stunted (HAZ < -2SD) | 83 | 0.55 ± 0.18 | 0.15 | 4.67 | <0.001 |
| | Not Stunted (HAZ ≥ -2SD) | 67 | 0.70 ± 0.21 | | | |
| Hemoglobin (g/dL) | Stunted (HAZ < -2SD) | 83 | 8.9 ± 1.1 | 0.9 | 3.98 | <0.001 |
| | Not Stunted (HAZ ≥ -2SD) | 67 | 9.8 ± 1.3 | | | |
| Serum Albumin (g/dL) | Stunted (HAZ < -2SD) | 83 | 2.9 ± 0.5 | 0.4 | 3.45 | 0.001 |
| | Not Stunted (HAZ ≥ -2SD) | 67 | 3.3 ± 0.6 | | | |

| Biomarker | Stunting Status | N | Mean ± SD | Mean Difference | t-statistic | p-value |
| --- | --- | --- | --- | --- | --- | --- |
|  | 2SD) |  |  |  |  |  |
| Serum Zinc ($\mu\text{g/dL}$) | Stunted (HAZ < -2SD) | 83 | 58.9 ± 7.2 | 5.2 | 4.12 | <0.001 |
| | Not Stunted (HAZ ≥ -2SD) | 67 | 64.1 ± 8.0 | | | |

Source: SPSS

### 3.3 Correlation Analysis Between Anthropometric Indices and Nutritional Biomarkers

Pearson’s correlation coefficients revealed consistently strong, statistically significant positive correlations between all anthropometric indices and nutritional biomarkers, and significant negative correlations with CRP (Table 3).

**Table 3:** Pearson’s Correlation Matrix Between Anthropometric Indices and Nutritional Biomarkers.

| Biomarker | Weight-for-Age Z-score (WAZ) | Height-for-Age Z-score (HAZ) | MUAC (cm) | Weight (kg) | Age (months) |
| --- | --- | --- | --- | --- | --- |
| Prealbumin | $r = 0.58^{**}$ | $r = 0.52^{**}$ | $r = 0.62^{**}$ | $r = 0.55^{**}$ | $r = 0.12$ |
| CRP | $r = -0.48^{**}$ | $r = -0.41^{**}$ | $r = -0.53^{**}$ | $r = -0.45^{**}$ | $r = -0.08$ |
| Serum Retinol | $r = 0.67^{**}$ | $r = 0.61^{**}$ | $r = 0.71^{**}$ | $r = 0.63^{**}$ | $r = 0.15$ |
| Hemoglobin | $r = 0.60^{**}$ | $r = 0.54^{**}$ | $r = 0.65^{**}$ | $r = 0.58^{**}$ | $r = 0.09$ |
| Serum Albumin | $r = 0.51^{**}$ | $r = 0.47^{**}$ | $r = 0.56^{**}$ | $r = 0.49^{**}$ | $r = 0.11$ |

| **Biomarker** | **Weight-for-Age Z-score (WAZ)** | **Height-for-Age Z-score (HAZ)** | **MUAC (cm)** | **Weight (kg)** | **Age (months)** |
| --- | --- | --- | --- | --- | --- |
| Serum Zinc | $r = 0.57^{**}$ | $r = 0.53^{**}$ | $r = 0.61^{**}$ | $r = 0.54^{**}$ | $r = 0.13$ |

Source: SPSS
**\*\*Correlation is significant at the 0.01 level (2-tailed).**

Table 3: The strongest correlations were observed between MUAC and nutritional biomarkers, with coefficients ranging from r = 0.56 (albumin) to r = 0.71 (serum retinol). Serum retinol showed the strongest correlations with all anthropometric indices, indicating that vitamin A status is particularly tightly linked to growth and nutritional status.

### 3.4 Prevalence of Biochemical Deficiencies by MUAC Category

Chi-square tests for trend revealed a striking dose-response relationship between the severity of acute malnutrition and the prevalence of all biochemical deficiencies (Table 4). For every biomarker assessed, the prevalence of deficiency increased progressively and significantly from normal to MAM to SAM categories (all p < 0.01).

**Table 4:**
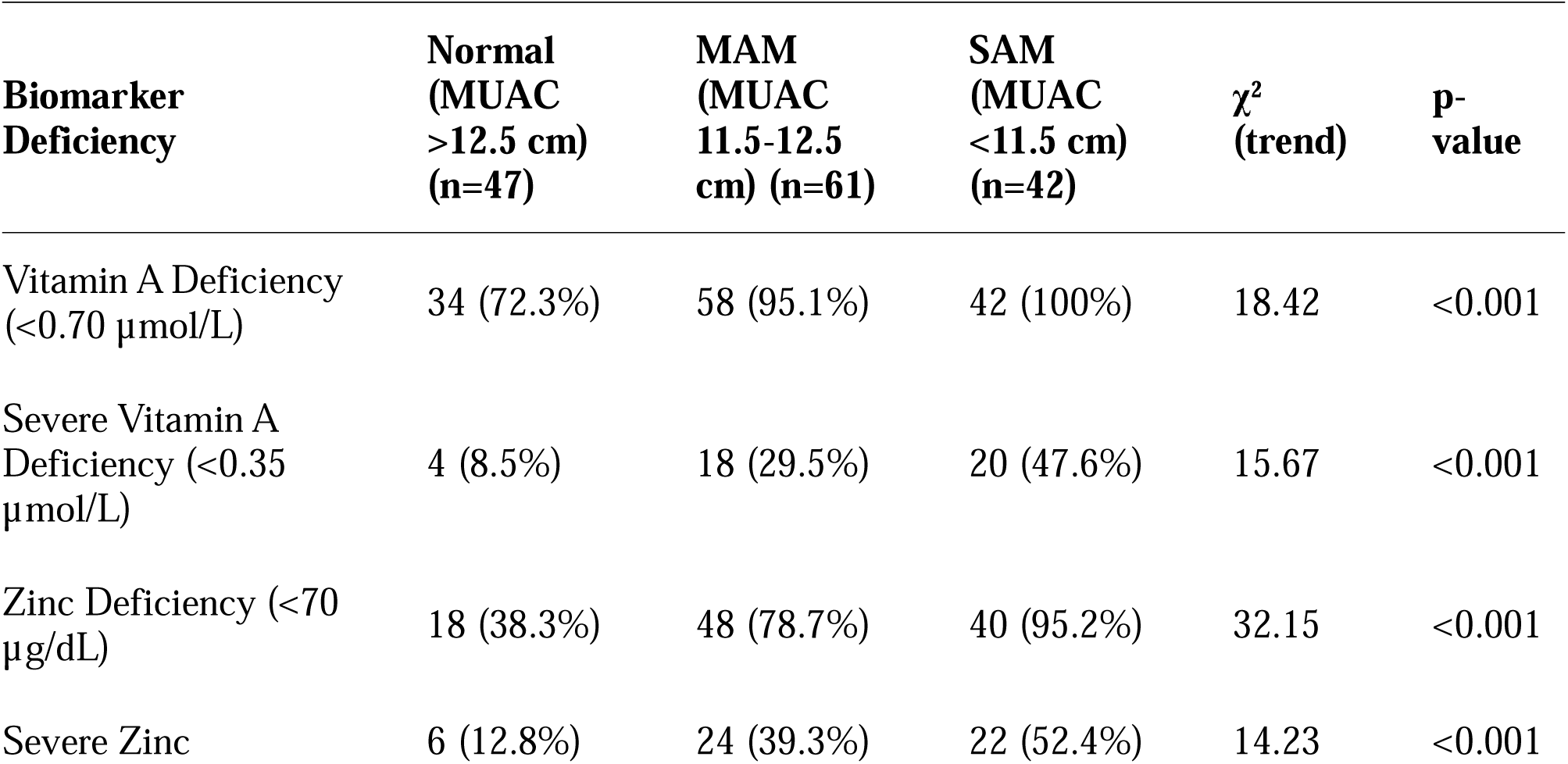
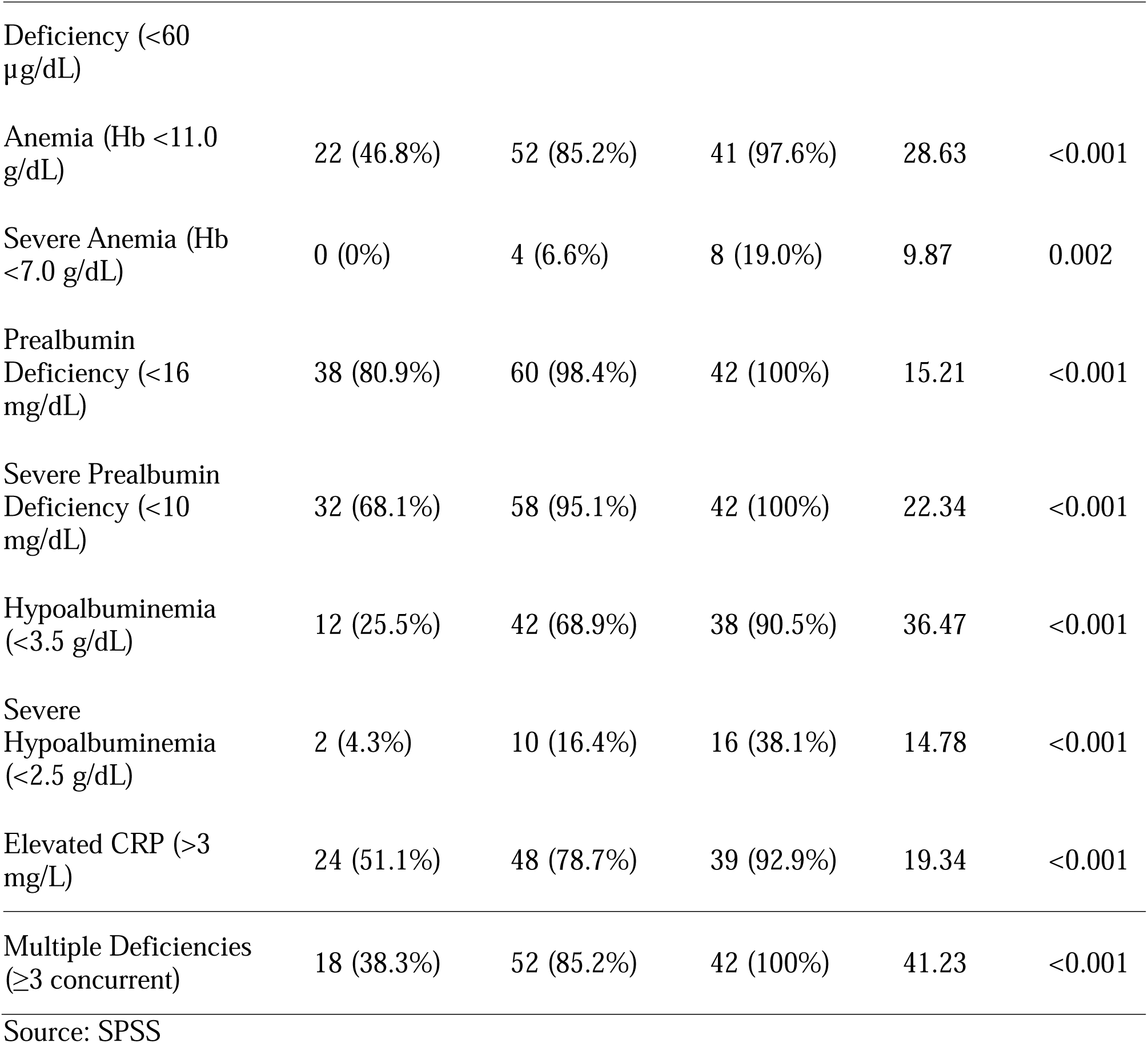
Prevalence of Biochemical Deficiencies by MUAC Category.

Table 4: Depicts the biochemical profile deteriorated markedly with worsening nutritional status. Among children with severe acute malnutrition (SAM), the burden of physiological depletion was near-universal: 100% exhibited vitamin A deficiency, prealbumin deficiency, and three or more concurrent micronutrient deficiencies; 97.6% were anemic; 95.2% were zinc deficient; 92.9% had elevated C-reactive protein (CRP); and 90.5% presented with hypoalbuminemia. Critically, even among children classified as “normal” by mid-upper arm circumference (MUAC), a substantial burden of hidden hunger was evident: 72.3% were vitamin A deficient, 46.8% were anemic, 38.3% were zinc deficient, 51.1% had elevated CRP, and 38.3% exhibited three or more concurrent deficiencies.

### 3.5 Logistic Regression: MUAC Category as Predictor of Biochemical Deficiencies

Binary logistic regression analysis quantified the dramatically increased risk of biochemical deficiencies associated with worsening anthropometric status (Table 5).

**Table 5:** Logistic Regression Analysis – MUAC Category as Predictor of Biochemical Deficiencies.

| Biochemical Deficiency | MUAC Category | Odds Ratio (OR) | 95% CI | p-value |
| --- | --- | --- | --- | --- |
| Vitamin A Deficiency | Normal | 1.00 (Reference) |  |  |
|  | MAM | 7.82 | 2.34 – 26.12 | 0.001 |
| | SAM | 18.42 | 4.18 – ∞ | <0.001 |
| Zinc Deficiency | Normal | 1.00 (Reference) |  |  |
|  | MAM | 5.94 | 2.61 – 13.52 | <0.001 |
|  | SAM | 28.44 | 6.12 – 132.18 | <0.001 |
| Anemia | Normal | 1.00 (Reference) |  |  |
|  | MAM | 6.68 | 2.78 – 16.05 | <0.001 |
|  | SAM | 42.68 | 5.38 – 338.41 | <0.001 |
| Prealbumin Deficiency | Normal | 1.00 (Reference) |  |  |
|  | MAM | 12.63 | 1.58 – 101.24 | 0.017 |
| | SAM | 22.84 | 1.32 – ∞ | 0.028 |
| Hypoalbuminemia | Normal | 1.00 (Reference) |  |  |
|  | MAM | 6.44 | 2.76 – 15.02 | <0.001 |
|  | SAM | 27.71 | 7.18 – 106.92 | <0.001 |
| Elevated CRP | Normal | 1.00 (Reference) |  |  |
|  | MAM | 3.54 | 1.58 – 7.92 | 0.002 |
|  | SAM | 12.19 | 3.38 – 43.95 | <0.001 |

Source: SPSS

Table 5: Shows that children with SAM had 18 times higher odds of vitamin A deficiency, 28 times higher odds of zinc deficiency, and 43 times higher odds of anemia compared to normal children. Even children with MAM had substantially elevated risks, with 6-13 times higher odds of deficiencies.

### 3.6 Discussion

This study provides robust statistical evidence for strong and clinically meaningful associations between anthropometric indices and biochemical markers of malnutrition among children under five in Sokoto State. The findings offer critical insights into the pathophysiology of childhood malnutrition, validate field screening tools, and carry profound implications for intervention design.

#### 3.6.1 Validation of Anthropometry and the Limits of Screening

The finding that children classified as malnourished by mid-upper arm circumference (MUAC) exhibit significantly lower levels of all nutritional biomarkers (p < 0.001) validates this field-friendly tool for identifying children at highest risk of physiological depletion. Mean prealbumin (5.8 mg/dL) and serum retinol (0.52 µmol/L) in malnourished children fell below clinical thresholds for severe deficiency, confirming active catabolism and the biochemical basis of observed growth failure (Ingenbleek & Young, 1994; Sommer & Vyas, 2012). Strong correlations between MUAC and all biomarkers (r = 0.56–0.71, p < 0.01) indicate that protein-energy reserves and micronutrient status deteriorate in parallel. However, these correlations, while robust, also demonstrate that anthropometry alone cannot fully capture the complexity of biochemical deficits, underscoring the need for integrated assessment approaches (Gibson, 2005).

#### 3.6.2 Biological Gradient: Severity of Wasting Reflects Depth of Depletion

A clear dose-response relationship was observed across MUAC categories: as acute malnutrition worsens, the prevalence and severity of biochemical deficiencies increase correspondingly. Among children with severe acute malnutrition (SAM), 100% exhibited vitamin A deficiency, prealbumin deficiency, and multiple concurrent deficiencies, while 97.6% were anemic and 95.2% were zinc deficient. These findings demonstrate unequivocally that SAM represents not merely a deficit of calories and protein, but a comprehensive metabolic failure affecting multiple physiological systems (Black et al., 2013).

Critically, even among children with moderate acute malnutrition (MAM), the burden of biochemical depletion was profound: 95.1% were vitamin A deficient, 85.2% anemic, and 78.7% zinc deficient. This finding has urgent programmatic implications. MAM is often viewed as a “mild” condition, but this study demonstrates that moderately wasted children harbor severe biochemical deficits that place them at high risk of deterioration to SAM and its associated mortality.

#### 3.6.3 Anthropometric Normalcy Does Not Equal Biochemical Sufficiency

Perhaps the most alarming finding is the high prevalence of biochemical deficiencies among children classified as “normal” by MUAC. Among these apparently well-nourished children, 72.3% were vitamin A deficient, 46.8% anemic, and 38.3% zinc deficient, with 38.3% exhibiting three or more concurrent deficiencies. This reveals a massive burden of “hidden hunger” that remains completely invisible to conventional anthropometric screening (Muthayya et al., 2013). The presence of elevated CRP in 51.1% of anthropometrically normal children further complicates the picture, indicating that even apparently well-nourished children experience systemic inflammation, likely from subclinical infections or Environmental Enteric Dysfunction (EED) associated with poor WASH conditions (Kosek et al., 2017). This inflammation may itself contribute to micronutrient deficiencies by depressing transport proteins and sequestering iron (Thurnham et al., 2021).

These findings have profound implications: reliance on anthropometry alone will miss the majority of children with micronutrient deficiencies; the true burden of malnutrition is substantially higher than indicated by anthropometric surveys alone; and interventions targeting only visibly malnourished children will fail to address the broader crisis of micronutrient deprivation affecting the entire pediatric population (Bailey et al., 2015).

#### 3.6.4 The Malnutrition-Inflammation Cycle

Significantly higher CRP levels in malnourished children (6.7 vs. 4.3 mg/L in MUAC comparison; 6.3 vs. 5.2 mg/L in stunting comparison) confirm that infection and inflammation are integral to the malnutrition syndrome. Negative correlations between CRP and all anthropometric indices (r = −0.41 to −0.53) demonstrate that as inflammation increases, nutritional status declines a pattern consistent with the classic malnutrition-infection cycle (Scrimshaw & SanGiovanni, 1997). The near-universal elevation of CRP in SAM children (92.9%) indicates that this cycle is actively operating in the most severely affected children and must be addressed integrally in treatment programs (Briend et al., 2021).

#### 3.6.5 Implications for Stunting

The finding that stunted children have significantly lower levels of all biomarkers, particularly vitamin A (0.55 vs. 0.70 µmol/L) and zinc (58.9 vs. 64.1 µg/dL), provides mechanistic insight into the etiology of linear growth failure. Vitamin A is essential for growth plate activity and bone remodeling, while zinc is critical for the growth hormone-IGF-1 axis and protein synthesis (Brown et al., 2022). Significantly higher CRP in stunted children supports the emerging consensus that chronic inflammation, particularly from EED, is a major contributor to stunting in low-income settings (Kosek et al., 2017).

#### 3.6.6 Programmatic Implications: Toward Integrated Interventions

The universal presence of multiple deficiencies in malnourished children (100% of SAM children had ≥3 deficiencies) demonstrates that malnutrition is not a series of single-nutrient problems but a comprehensive metabolic failure. This argues forcefully for multi-micronutrient interventions rather than single-nutrient supplementation (Bhutta et al., 2020). The use of multiple micronutrient powders (MNPs) for home fortification of complementary foods should be considered for all children 6–59 months in this population.

Elevated CRP confirms that nutritional rehabilitation must be integrated with infection treatment and prevention, including routine deworming, malaria chemoprevention, promotion of timely vaccination, and crucially, improvements in water, sanitation, and hygiene (WASH) to reduce pathogen exposure driving EED (Humphrey, 2009). Attempting to treat malnutrition without addressing its infectious and environmental drivers is unlikely to achieve sustained success.

The high prevalence of biochemical deficiencies among anthropometrically normal children argues for population-level interventions rather than targeting only visibly malnourished children. Universal supplementation strategies (e.g., twice-yearly vitamin A campaigns, multiple micronutrient powders for all young children) are justified in this context, as the entire pediatric population is at risk of hidden hunger (Imdad et al., 2017). Food-based approaches that improve overall dietary diversity and quality including promotion of home gardening, small-scale livestock production, and nutrition education remain essential long-term strategies for addressing the root causes of multiple, concurrent deficiencies (Gibson et al., 2018).

## 4. Conclusion and Recommendations

This study provides robust evidence that anthropometric indicators, particularly MUAC, are valid proxies for identifying children at highest risk of biochemical depletion. The strong, consistent, and dose-response relationships between wasting severity and biomarker deficiencies validate current screening protocols that use MUAC to identify children needing therapeutic feeding. Children with SAM have near-universal, severe, and multiple biochemical deficiencies requiring comprehensive, multi-micronutrient rehabilitation integrated with infection treatment.

However, the high prevalence of “hidden hunger” among anthropometrically normal children reveals a critical limitation of sole reliance on anthropometry. Most children with micronutrient deficiencies in this population are not visibly wasted and would be missed by conventional screening. This argues for integrated assessment approaches that combine anthropometry with biochemical monitoring at sentinel sites, and for population-level interventions that address the full spectrum of malnutrition from visible wasting to invisible biochemical deficiencies. Based on these findings, the following recommendations are made:

i. Maintain and Strengthen Anthropometric Screening: Current protocols using MUAC to identify children with MAM and SAM for treatment should be maintained and strengthened, as these children have profound biochemical depletion requiring urgent intervention.
ii. Integrate Multi-Micronutrient Interventions: Given the universal presence of multiple deficiencies in malnourished children, treatment protocols should provide comprehensive multi-micronutrient supplementation rather than focusing solely on energy and protein. Ready-to-Use Therapeutic Foods (RUTF) already contain multiple micronutrients, but this finding justifies their continued use and expansion.
iii. Implement Population-Level Micronutrient Strategies: The high burden of deficiencies among anthropometrically normal children justifies universal interventions, including twice-yearly high-dose vitamin A supplementation, multiple micronutrient powders (MNPs) for children 6-23 months, and consideration of preventive zinc supplementation.
iv. Integrate Nutrition and Infection Control: The elevated CRP in malnourished children confirms that nutritional interventions must be integrated with infection treatment and prevention, including deworming, malaria control, immunization, and WASH improvements.
v. Enhance Surveillance with Biomarkers: While routine biomarker assessment may not be feasible at all levels, sentinel surveillance sites should integrate biochemical monitoring to track the true burden of “hidden hunger” and evaluate the impact of interventions.
vi. Address Dietary Diversity as a Long-Term Solution: The multiple, concurrent deficiencies documented in this study reflect a fundamental failure of dietary quality. Long-term strategies must focus on improving dietary diversity through nutrition education, agricultural interventions (home gardening, biofortified crops, small-scale livestock), and social protection programs that increase household access to nutrient-rich foods.

The anthropometric-biochemical link established in this study closes the logical loop of this investigation: from the visible manifestation of malnutrition through its biochemical underpinnings to the physiological consequences of inflammation and metabolic dysregulation. This integrated understanding provides the evidence base for comprehensive, multi-sectoral interventions to address one of the most severe child nutrition crises documented in contemporary Nigerian public health research.

### 4.1 Strengths and Limitations

The primary strength of this study is its integration of comprehensive anthropometric assessment with a robust panel of biochemical markers, allowing for a nuanced understanding of the relationship between visible and invisible dimensions of malnutrition. The use of standardized measurement techniques, validated laboratory methods, and internationally recognized classification thresholds enhances the reliability and comparability of the findings. The multiple analytical approaches t-tests, correlation, chi-square trend, and logistic regression provide complementary perspectives on the anthropometric-biochemical relationship.

Several limitations should be acknowledged. The cross-sectional design captures associations at a single point in time and cannot establish causality or temporal sequence. The sample was drawn from PHC attendees, which may introduce selection bias toward children already in contact with the health system; this could either overestimate the burden (if sicker children attend PHCs) or underestimate it (if the most vulnerable never access care). The relatively small sample size, while adequate for detecting the large effect sizes observed, limits the precision of some estimates, particularly in logistic regression where confidence intervals were wide for some parameters due to near-complete separation. Finally, we did not assess all potential confounders (e.g., malaria parasitemia, helminth infection, socioeconomic status in multivariate models), which could provide a more complete understanding of the pathways linking anthropometric and biochemical status.

## Data Availability

All data produced in the present work are contained in the manuscript

